# Poor odor identification predicts mortality risk in older adults without neurodegenerative diseases: the Shanghai Aging Study

**DOI:** 10.1101/2020.11.10.20229518

**Authors:** Zhenxu Xiao, Qianhua Zhao, Xiaoniu Liang, Wanqing Wu, Yang Cao, Ding Ding

## Abstract

**Background:** Irrespective of neurodegeneration, the decline of olfactory function might also reflect a wider range of pathological conditions contributing to mortality. However, the potential explanations and their predictive values have rarely been reported.

**Methods:** A total of 1,433 older adults aged ≥ 60 years without neurodegenerative disease were administered a follow-up of 8.6 years on average. Sniffin’ Sticks Screening Test was used to assess the olfactory identification at baseline. Survival status during the follow-up was obtained from the local Center for Disease Control and Prevention. Bidirectional stepwise Cox proportional hazards regression was used to identify variables associated with mortality. Two predictive models were constructed by statistical learning methods.

**Results:** All-cause mortality rate was 1.1/100 person-years during the follow-up. Sex (HR = 0.65, 95%CI 0.46 - 0.93), age (HR = 1.12, 95%CI 1.10 - 1.15), chronic kidney disease (HR = 1.88, 95%CI 1.09 - 3.25), low density lipoprotein (HR = 0.81, 95%CI 0.67 - 0.99), anemia (HR = 2.74, 95%CI 1.19 - 6.30), and fail to identify coffee odor (HR = 1.96, 95%CI 1.19 - 3.23) were significantly associated with all-cause mortality. The logistic regression and the random forest predictive models showed similar predictive accuracy (0.91 and 0.90) and area under the receiver operating characteristic curve (0.77 and 0.75).

**Conclusions:** The association between poor olfactory and long-term mortality was verified among Chinese older population. Certain odors identification ability may contribute to the prediction of long-term mortality along with other important risk factors.

## Introduction

Olfaction plays a crucial role in human health by influencing dietary behavior, increasing the awareness of environmental risks, and affecting social interaction (1). Olfactory impairment is an age-related disorder (2). Previous studies have indicated the potential role of olfactory impairment as a marker of underlying physiologic processes or pathology associating with aging and reduced survival in older adults.

Olfactory impairment is regarded as an early marker of impending neurodegenerative diseases, such as Alzheimer’s disease (AD), mild cognitive impairment (MCI), Parkinson’s disease (PD), dementia with Lewy bodies, frontotemporal lobar degeneration, and Huntington’s disease (3), which in turn is related to greater mortality risk (4). An Australian cohort study concluded that the relationship between olfaction and mortality was mediated by cognitive impairment in older adults (5). However, neurodegenerative diseases were reported to contribute to part of the mortality risk in other cohort studies. The Washington Heights/Inwood Columbia Aging Project found a persistent association between odor identification and future mortality in older adults, independent of prospective dementia diagnoses (6). The Betula Study in Sweden indicated that dementia does not attenuate the association between olfactory loss and mortality (7). The Epidemiology of Hearing Loss Study (EHLS) in Wisconsin also found a significant association after adjusting cognitive impairment and other potential confounders (8). The Health, Aging, and Body Composition (Health ABC) study found that poor olfaction was associated with higher long-term mortality, particularly among older adults with excellent to good health at baseline. The study also suggests that impaired olfaction is more than a marker of poor overall health because only one-third of the poor olfaction associated mortality could be explained by dementia or Parkinson’s disease and weight loss (9). Therefore, irrespective of neurodegeneration, the decline of olfactory function might also reflect a wider range of pathological conditions (10), declining cell regeneration (11), and age-related accumulation of environmental exposures (12), which could contribute to mortality. However, empirical data of potential explanations and their predictive values have rarely been reported.

This study aimed to verify the hypothesis that poor olfactory identification predicts long-term mortality risk in older adults without neurodegenerative diseases by analyzing the longitudinal data of the Shanghai Aging Study. We also evaluated the importance of specific odor identification and other known risk factors for predicting mortality and the predictive ability of models constructed by statistical learning methods.

## Methods

### Study participants

The Shanghai Aging Study is a population-based prospective cohort study aiming to investigate the prevalence, incidence, and risk factors for dementia and cognitive impairment among older adults residing in a community of downtown Shanghai, China. The detailed study design and recruitment process of the cohort have been described elsewhere (13).

In the current study, participants were older adults aged 60 years or older who completed the olfactory identification test at the baseline (2009-2010). Participants who met the following criteria were excluded: 1) with dementia, MCI, PD, or other neurodegenerative diseases; 2) undergone maxillofacial surgery, with lesions of the nose or the paranasal sinuses (e.g. rhinosinusitis, polyposis, and allergic rhinitis); 3) with chronic obstructive pulmonary disease, asthma, acute upper respiratory tract infection within 1 week, or alcohol and drug abuse, which may affect the test of olfactory function (14); 4) had mental retardation or schizophrenia confirmed on their medical record; 5) had severe problems of vision, hearing, or speaking, and were not able to participate actively in the neuropsychological evaluation.

This study was approved by the Medical Ethical Committee of Huashan Hospital, Fudan University, Shanghai, China (approval number: 2009-195). All participants and/or their legal guardians have signed written informed consent for participation in the study.

### Demographics, lifestyle, and medical history

The demographic and lifestyle characteristics of the participants, including age, gender, years of education, cigarette smoking, alcohol consumption, physical activity, and the activity of daily living (ADL) were collected via interviewer-administered questionnaires (13, 15). The height and weight of each participant were measured to calculate Body Mass Index (BMI). Medical histories of the participants, including physician-diagnosed hypertension, diabetes, stroke, coronary heart disease, depression, cancer, chronic kidney disease, anemia, urinary tract infections, and chronic bronchitis were confirmed from the medical records (13, 15).

### Olfactory identification test

The olfactory identification was assessed using the Sniffin’ Sticks Screening Test 12 (SSST-12), which was produced by the Burghart Medical Technology (16). The 12 validated common odors (orange, leather, cinnamon, peppermint, banana, lemon, liquorice, coffee, cloves, pineapple, rose, and fish) were presented in 12 different felt-tip sticks (17). Participants were asked to sniff each odor and to name it with or without the help of the choices. The procedure of administrating the SSST-12 was detailed described elsewhere (14).

### Laboratory tests

Fasting blood samples were drawn by a research nurse to test total cholesterol (TC), triglycerides (TG), low density lipoprotein (LDL), and high-density lipoprotein (HDL) of study participants. Apolipoprotein (APOE) genotyping was conducted by the TaqmanSNP method based on the blood or saliva samples (18). The presence of at least one ε4 allele was regarded as APOE-ε4 allele positive.

### Diagnosis of neurodegenerative diseases

We excluded participants with a diagnosis of neurodegenerative diseases either confirmed by medical history or diagnosed at the clinical interview at the baseline. Neurologists from Huashan Hospital conducted neurologic examinations assessing the sensory neurons, motor responses, and reflexes of each participant. Medical histories of neurodegenerative diseases, e.g. Dementia, AD, MCI, PD, frontotemporal lobar degeneration, and Huntington’s disease were reported by participants and confirmed from their medical records. Additionally, a battery of neuropsychological tests covering domains of global cognition, executive function, spatial construction function, memory, language, and attention was used to assess the cognitive function of the participants. The test battery contains the Mini-mental State Examination (MMSE) (19), Conflicting Instructions Task (Go/No Go Task), Stick Test, Modified Common Objects Sorting Test, Auditory Verbal Learning Test, Modified Fuld Object Memory Evaluation, Trail-making test A&B, and Ren Min Bi (Chinese currency) test. The elaborate description and application method of these tests were reported elsewhere (20, 21). A panel of experts reached a consensus diagnosis of dementia or non-dementia based on the criteria of Diagnostic and Statistical Manual of Mental Disorders, the 4th edition (DSM-V) (22). MCI was diagnosed based on the Peterson criteria (23).

### Follow-up and mortality surveillance

Study participants were prospectively followed up after baseline clinical interviews. Survival status of participants from baseline to December 31, 2019 was obtained from the mortality surveillance system of the local Center for Disease Control and Prevention, which is responsible for verifying the date of death and causes of death from the death certificate (24).

### Statistical analysis

The continuous variables were presented as mean with standard deviation (SD), and the categorical variables were presented as count and percentage (%). The mortality rate was calculated as the number of dead participants divided by total person-years of follow-up. The participants were divided into good (SSST-12 score > 8) or poor (SSST-12 score ≤ 8) olfactory identification function by the median of the SSST-12 scores. The Pearson’s chi-squared test was used to test the differences between groups for categorical variables, and the Student’s t-test and Mann-Whitney U test were used for continuous variables. The Kaplan-Meier curve and the Log-rank test were used to estimate and compare the mortality rates of participants with good and poor olfactory identification.

The correlation between two continuous variables was evaluated using the Pearson’s correlation coefficient. The point-biserial correlation coefficient was used between a binary variable and a continuous variable (25), and the phi coefficient was used between two binary variables (26). Heatmap was used to visualize the multicollinearity among independent variables, e.g. LDL, TC, olfactory sum score, and each specific odor (eFigure 1). Two-tailed P-values < 0.05 were considered as statistically significant.

The bidirectional stepwise Cox proportional hazards regression analysis (p for entry = 0.05, p for remove = 0.10) was used to identify variables associated with mortality among the independent variables, including demographics, odors, lifestyles, chronic diseases, and laboratory indexes at baseline shown in Table 1.

**Table 1.** Baseline demographic characteristics of study participants with poor and good olfactory identification.

| Baseline characteristics | All<br>(N = 1,433) | Olfactory identification |  | *P*-value* |
| --- | --- | --- | --- | --- |
|  |  | Poor<br>SSST-12<br>Score ≤8<br>(N = 768) | Good<br>SSST-12<br>Score >8<br>(N = 665) |  |
| Gender, female, n (%) | 777 (54.2) | 398 (51.8) | 379 (57.0) | 0.050 |
| Age, years, mean (SD) | 68.9 (6.8) | 70.2 (7.0) | 67.4 (6.3) | <0.001 |
| Education, years, mean (SD) | 12.8 (3.5) | 12.5 (3.8) | 13.1 (3.1) | <0.001 |
| Body mass index, mean (SD) | 24.3 (3.4) | 24.3 (3.5) | 24.2 (3.4) | 0.342 |
| Cigarette smoking, n (%) | 141 (9.8) | 83 (10.8) | 58 (8.7) | 0.186 |
| Alcohol drinking, n (%) | 114 (8.0) | 67 (8.7) | 47 (7.1) | 0.248 |
| Physical activity, mean (SD) | 27.2 (25.1) | 28.0 (26.2) | 26.2 (23.7) | 0.161 |
| Coronary heart disease, n (%) | 149 (10.4) | 88 (11.5) | 61 (9.2) | 0.158 |
| Hypertension, n (%) | 756 (52.8) | 422 (54.9) | 334 (50.2) | 0.074 |
| Diabetes, n (%) | 179 (12.5) | 97 (12.6) | 82 (12.3) | 0.864 |
| Depression, n (%) | 205 (14.3) | 109 (14.2) | 96 (14.4) | 0.896 |
| Stroke, n (%) | 153 (10.7) | 93 (12.1) | 60 (9.0) | 0.059 |
| Cancer, n (%) | 154 (10.7) | 75 (9.8) | 79 (11.9) | 0.198 |
| Chronic kidney disease, n (%) | 80 (5.6) | 42 (5.5) | 38 (5.7) | 0.840 |
| Anemia, n (%) | 23 (1.6) | 11 (1.4) | 12 (1.8) | 0.576 |
| Urinary tract infections, n (%) | 453 (31.6) | 246 (32.0) | 207 (31.1) | 0.714 |
| Chronic bronchitis, n (%) | 203 (14.2) | 136 (17.7) | 67 (10.1) | <0.001 |
| MMSE score, mean (SD) | 28.7 (1.5) | 28.5 (1.6) | 28.9 (1.3) | <0.001 |
| ADL scores, mean (SD) | 20.2 (1.9) | 20.3 (1.9) | 20.2 (1.8) | 0.366 |
| Serum total cholesterol, mean (SD) | 5.4 (1.1) | 5.4 (1.1) | 5.5 (1.1) | 0.011 |
| Serum triglyceride, mean (SD) | 1.7 (1.0) | 1.7 (1.1) | 1.8 (1.0) | 0.188 |
| Serum high-density lipoprotein, mean (SD) | 1.3 (0.3) | 1.3 (0.4) | 1.3 (0.3) | 0.573 |
| Serum low density lipoprotein, mean (SD) | 3.3 (0.9) | 3.2 (0.9) | 3.3 (0.9) | 0.095 |
| Apolipoprotein E ε4 positive, n (%) | 251 (17.9) | 132 (17.6) | 119 (18.3) | 0.731 |
| SSST-12 score, mean (SD) | 8.2 (1.9) | 6.8 (1.5) | 9.8 (0.9) | <0.001 |
| Orange<sup>a</sup>, n (%) | 1,120 (78.2) | 527 (68.6) | 593 (89.2) | <0.001 |
| Leather<sup>a</sup>, n (%) | 842 (58.8) | 330 (43.0) | 512 (77.0) | <0.001 |
| Cinnamon<sup>a</sup>, n (%) | 647 (45.2) | 244 (31.8) | 403 (60.6) | <0.001 |
| Peppermint<sup>a</sup>, n (%) | 1319 (92.0) | 670 (87.2) | 649 (97.6) | <0.001 |
| Banana<sup>a</sup>, n (%) | 974 (68.0) | 406 (52.9) | 568 (85.4) | <0.001 |
| Lemon<sup>a</sup>, n (%) | 787 (54.9) | 343 (44.7) | 444 (66.8) | <0.001 |
| Liquorice<sup>a</sup>, n (%) | 806 (56.2) | 309 (40.2) | 497 (74.7) | <0.001 |
| Coffee<sup>a</sup>, n (%) | 1343 (93.7) | 684 (89.1) | 659 (99.1) | <0.001 |
| Cloves<sup>a</sup>, n (%) | 747 (52.1) | 306 (39.8) | 441 (66.3) | <0.001 |
| Pineapple<sup>a</sup>, n (%) | 1012 (70.6) | 429 (55.9) | 583 (87.7) | <0.001 |
| Rose<sup>a</sup>, n (%) | 937 (65.4) | 383 (49.9) | 554 (83.3) | <0.001 |
| Fish<sup>a</sup>, n (%) | 1214 (84.7) | 586 (76.3) | 628 (94.4) | <0.001 |

Note: SSST-12, Sniffin' Sticks Screening Test 12; SD, standard deviation; MMSE, Mini-mental State Examination; ADL, Activities of Daily Living.
a, 'n' represents the number of participants who were able to identify this specific odor.
\* Compared between the participants with poor and good olfactory identification function.

Variables’ importance was evaluated using both the permutation importance (PI) by the multivariate logistic regression (LR) model and Gini importance (GI) by the random forest (RF) model (27-29) (eMethods 1). We used the K-fold cross-validation method to calculate the predictive ability indices (30) (eMethods 2), including accuracy, sensitivity, specificity, and area under the receiver operating characteristic (ROC) curve (31). The area under the ROC curve (AUC) of a predictive model greater than 0.7 was defined as acceptable prediction ability (32, 33).

The descriptive analyses and Cox proportional hazards regression model were conducted in Stata 16.1 (StataCorp LLC, College Station, TX, USA). PI and GI evaluation by the LR and RF models were achieved in Python 3.6 (Python Software Foundation, https://www.python.org/) using packages scikit-learn 0.22.1 (34) and ELI5 0.10.1 (35). All the computation was operated on a computer with the 64-bit Windows 7 Enterprise operating system (Service Pack 1), Intel ® Core TM i5-4210U CPU of 2.40 GHz, and 16.0 GB installed random access memory.

## Results

Baseline characteristics of the 1,433 enrolled participants were presented in Table 1. In general, participants with poor olfactory identification (n = 768) were older (70.2 vs. 67.4 years, *P* < 0.001), received less education (12.5 *vs*. 13.1 years, *P* < 0.001), and had lower MMSE scores (28.5 *vs*. 28.9, *P* < 0.001) comparing with participants with good olfactory identification (n = 665). Participants with poor olfactory identification were more vulnerable to chronic bronchitis (17.7 % *vs*. 10.1 %, *P* < 0.001) and having lower TC (5.4 *vs*. 5.5, *P* = 0.011). The total SSST-12 score and proportions of correctly identifying each odor were significantly different between participants with good and poor olfactory identification (all *P* < 0.001).

During the follow-up of averagely 8.6 (SD = 1.4) years, the all-cause mortality rate was 1.1/100 person-years in total participants. Participants with poor olfactory identification had a significantly higher mortality rate than those with good olfactory identification (1.3/100 person-years *vs*. 0.8/100 person-years, *P* = 0.004). The Kaplan-Meier curve in Figure 1 presented the cumulated mortality of the participants with good or poor olfactory identification.

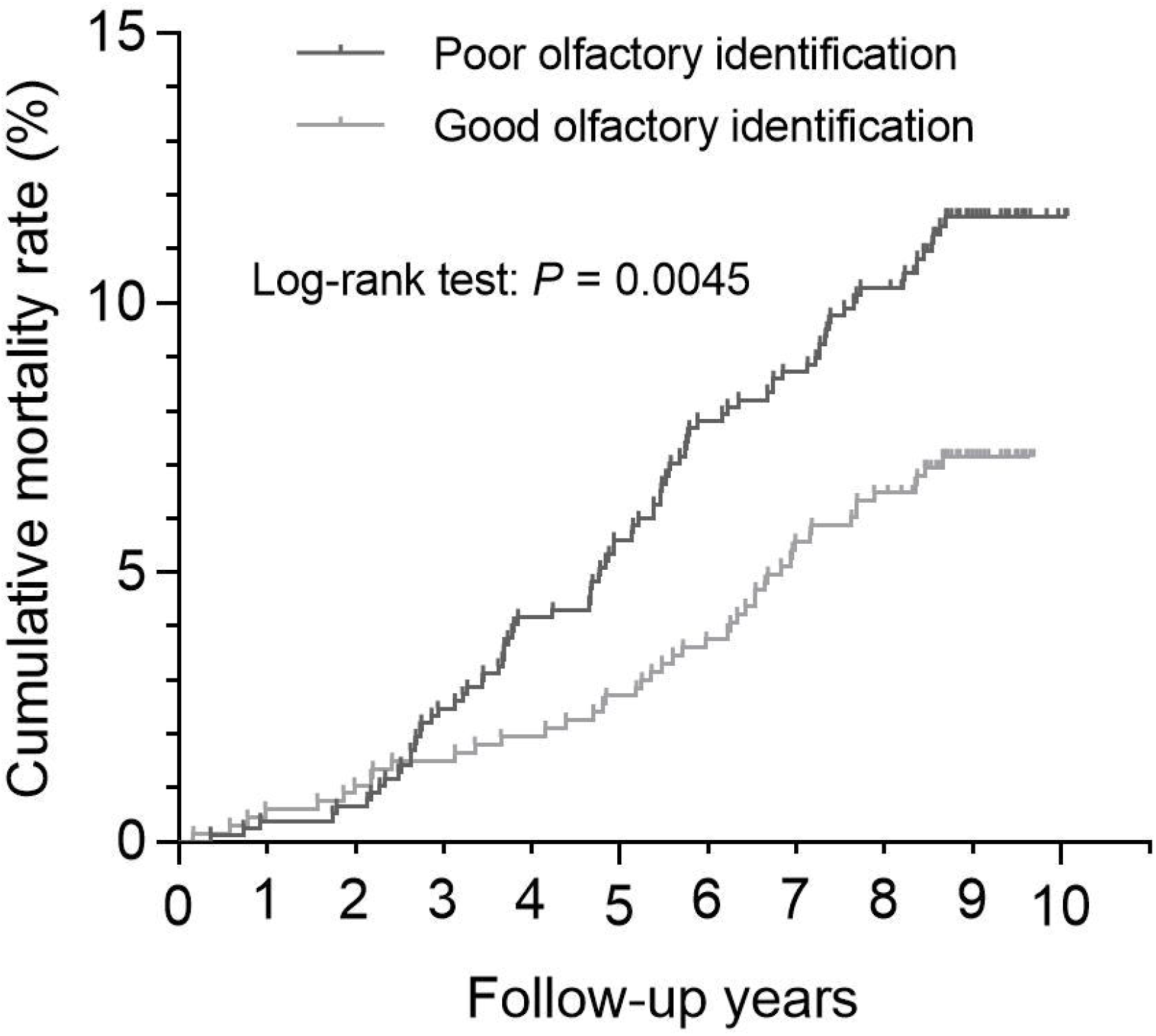

The bidirectional stepwise Cox proportional hazards regression analysis identified that sex (HR = 0.65, 95%CI 0.46 - 0.93, *P* = 0.019), age (HR = 1.12, 95%CI 1.10 - 1.15, *P* < 0.001), chronic kidney disease (HR = 1.88, 95%CI 1.09 - 3.25, *P* = 0.024), LDL (HR = 0.81, 95%CI 0.67 - 0.99, *P* = 0.043), anemia (HR = 2.74, 95%CI 1.19 - 6.30, *P* = 0.017), and fail to identify coffee odor (HR = 1.96, 95%CI 1.19 - 3.23, *P* = 0.009) were significantly associated with all-cause mortality (Table 2). PI and GI by LR and RF models indicated the similar order of the identified important variables, i.e. age, ADL, LDL, chronic kidney disease, fail to identify coffee, anemia, sex, fail to identify rose, and fish (Figure 2).

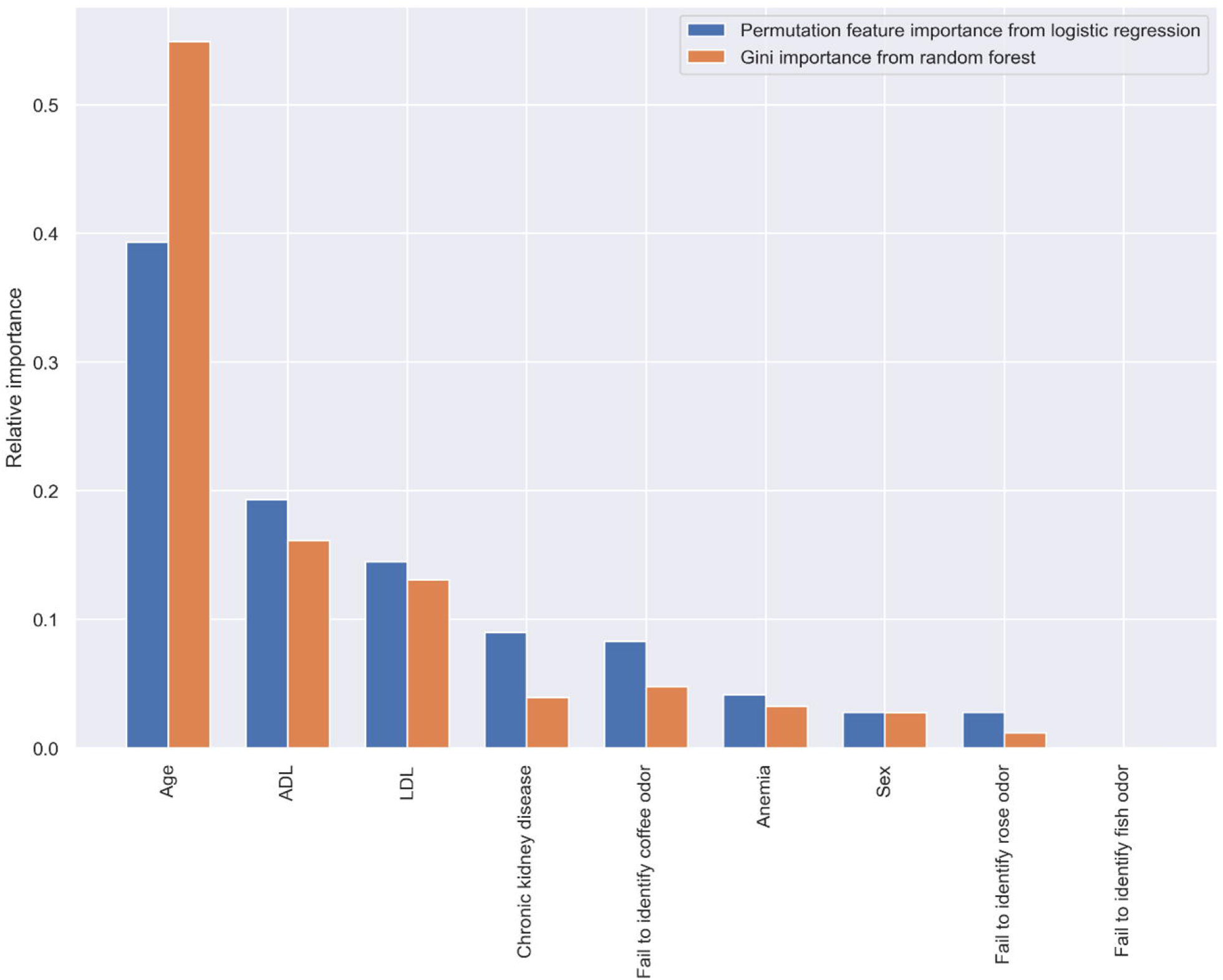

**Table 2.** Hazard ratios (95%CI) of the selected variables associated with all-cause mortality by Stepwise Cox regression analysis

|  | HR | 95% CI | *P*-value |
| --- | --- | --- | --- |
| Sex, female | 0.65 | 0.46 - 0.93 | 0.019 |
| Age | 1.12 | 1.10 - 1.15 | <0.001 |
| Failing to identify Rose odor | 1.38 | 0.97 - 1.96 | 0.074 |
| ADL score | 1.05 | 1.00 - 1.10 | 0.053 |
| Chronic kidney disease | 1.88 | 1.09 - 3.25 | 0.024 |
| Serum low density lipoprotein | 0.81 | 0.67 - 0.99 | 0.043 |
| Failing to identify Fish odor | 0.61 | 0.37 - 1.01 | 0.053 |
| Anemia | 2.74 | 1.19 - 6.30 | 0.017 |
| Failing to identify Coffee odor | 1.96 | 1.19 - 3.23 | 0.009 |

Note: HR, hazard ratio; CI, confidence interval; ADL, Activity of Daily Living Scale.

Figure 3 exhibited the predictive ability indices of the LR and RF models constructed using the selected important variables. The LR and RF models showed similar predictive ability with AUC = 0.77 and 0.75, and accuracy = 0.91 and 0.90.

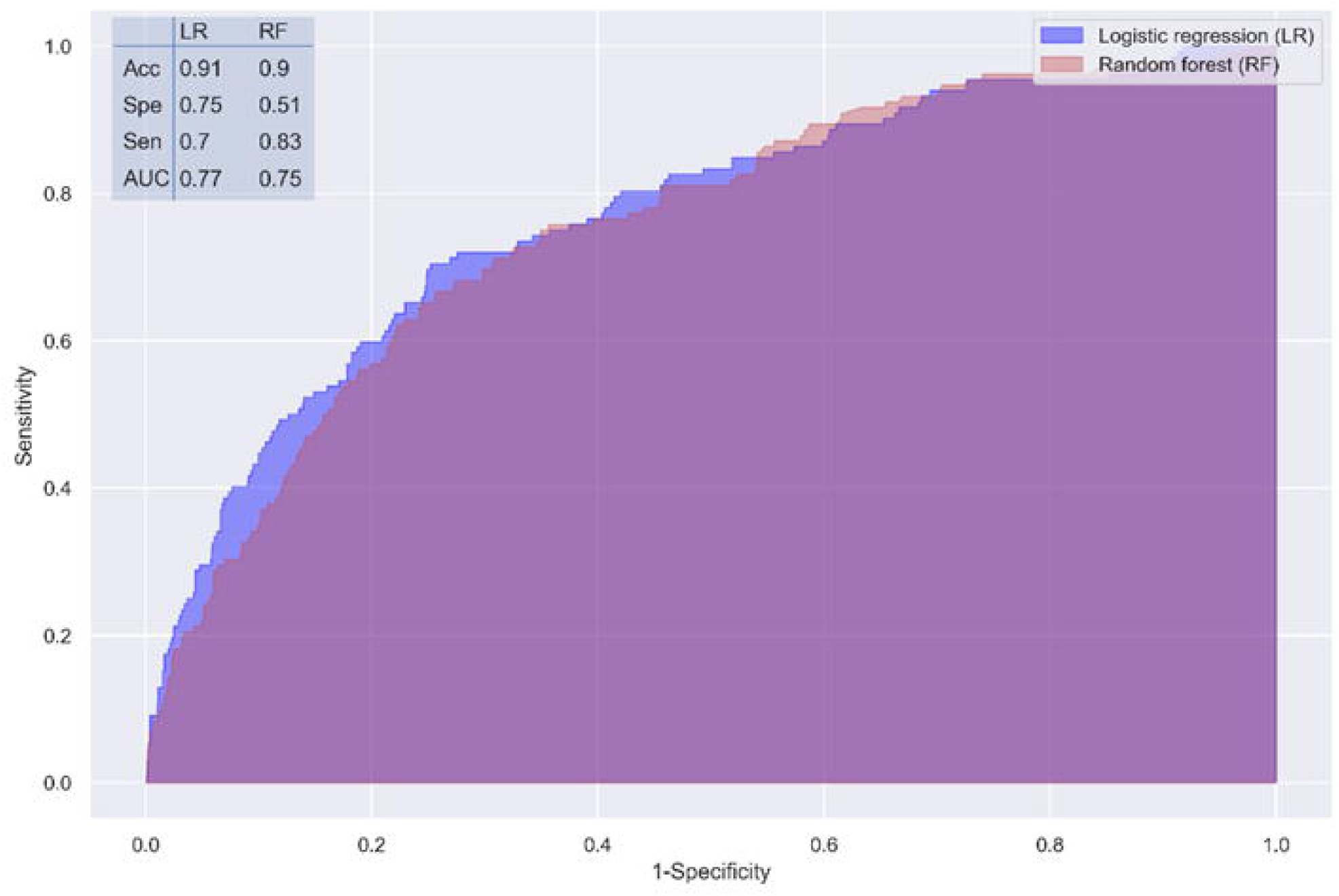

## Discussion

The current study verified the association between poor olfactory identification and long-term mortality risk in older adults without neurodegenerative diseases. Prediction models containing nine important variables showed acceptable predictive abilities in predicting all-cause mortality. Specifically, the ability to identify coffee odor showed unique importance on the risk of death.

Many population-based longitudinal studies have reported the association between olfactory impairment and the mortality risk in older adults (4-10, 36, 37). Although the olfactory tests and the covariates were diverse, all the studies concluded consistent results that participants with poor olfactory identification had significantly higher mortality risk. Olfactory impairment is considered as the preclinical symptom of neurodegenerative diseases, which may play a key role in this relationship. However, several previous studies indicated that neurodegenerative diseases only could explain part of the association of olfactory impairment with future mortality. In the Health ABC Study, people with poor olfactory function were 1.46 and 1.30 times more likely to die during 10 follow-up years and 13 follow-up years, while neurodegenerative diseases explained 22% higher mortality in 10 years (9). The Blue Mountains Eye Study demonstrated the association between the moderate olfactory loss and the 68% increased risk of five-year all-cause mortality disappeared when adjusting for cognitive impairment (5). Our results demonstrated that in those older adults without neurodegenerative disease, even MCI, poor olfaction could still be an essential contributor in predicting long-term mortality.

Previous studies only demonstrated the association of olfactory function and mortality risk. However, to our knowledge, the predictive value assessed by statistical learning models has not been reported. Abundant demographic and medical historical variables collected in our Shanghai Aging Study offered us a unique opportunity to screen the valuable variables for the predictive models. The permuting method took both the main effect of a variable and the mutual effect with other variables into consideration in prediction models. In the LR model, PI was measured by looking at how much the accuracy decreases when the variable’s information is not available (38). In the RF model, GI was calculated as the sum over the number of splits (across all trees) that include the variable, proportionally to the number of samples in each split (27). We used the two predictive models with two indexes and showed much similar AUCs, which could be verified mutually and improve the robustness of our results.

To date, several possible mechanisms have explained the association between olfactory impairment and the risk of death, regardless of the impact of neurodegenerative diseases. First, because the olfactory function partly determines the eating experience, poor olfaction could induce a poor appetite, resulting low body weight, low BMI, or even malnutrition (39, 40). These consequences and the contribution to death have already been confirmed in several epidemiological studies (5, 9, 10). Second, olfactory impairment might be a manifestation of the declining ability of neurogenesis and plasticity. Since adult neurogenesis exists in the olfactory system peripherally and centrally, olfactory impairment could be regarded as a sign of brain aging (41). Third, olfactory impairment was also found associated with cardiovascular diseases and diabetes mellitus (DM) (5, 9). The Health ABC study concluded that poor olfaction was modestly associated with death from cardiovascular disease (9). Gouveri et al. found that patients with type 2 diabetes mellitus (T2DM) with diabetic retinopathy had significantly worse olfactory function than T2DM patients without complication (42). It indicated that microvascular injury might play a role in olfactory impairment among T2DM patients. Fourth, some environmental exposures, such as pollution and toxins, could directly cause central nervous system diseases through the olfactory nerve or indirectly cause pulmonary and cardiovascular diseases, which induced higher mortality risk (43, 44). This so-called *olfactory vector hypothesis* emphasized that some people with PD, which present with smell loss concomitantly, may be caused by environmental agents that enter the brain via olfactory mucosa (12). Lastly, age-related hyposmia could manifest regenerative dysfunction and atrophic of the olfactory epithelium, which may be caused by telomere shortening (11).

Although the underlying mechanism of the relationship between coffee identification and death has not been illuminated, there are still some clues. Our previous study demonstrated the inability to smell coffee was related to the annual decline rate of the MMSE scores (45). An interesting study found that recognition of the coffee odor was faster in caffeine consumers than non-consumers, and high-caffeine consumers had greater olfactory sensitivity for the coffee odor than the moderate- or non-consumers (46). According to these results, we suspect that people who could not identify coffee were less likely to drink coffee. Since many large-sampled studies found that coffee drinkers were inversely associated with total and cause-specific mortality (47, 48), failing to smell coffee may be associated with higher mortality. Whether there are other mechanisms between coffee identification and death should be further explored.

There were some limitations in this study. First, our prediction models were constructed based on Han Chinese older adults aged ≥ 60 years living in an urban community, which limited the generalization of results from the current study. Second, although we excluded people with neurodegenerative diseases based on medical records and examinations of cognitive impairment, there still might be some participants at the preclinical phase of neurodegenerative diseases that could not be detected. Third, the SSST-12 test only covers 12 common odors. There might be other odors that could have predictive value to predict mortality. More odors which are crucial in our daily lives and their potential value need to be further explored by using olfactory tests with more odor samples. Finally, better performance of predictive models depends on more observation data. The relatively small sample size in our study limited us to construct prediction models of cause-specific mortality further.

In conclusion, the association between poor olfactory and long-term mortality was verified among the Chinese older adults without neurodegenerative diseases. Certain odors identification ability may contribute to the prediction of long-term mortality along with other important risk factors.

## Supplementary Material

### eMethods 1. The description of PI in LR model and GI in RF model

In the LR model, the PI was calculated for each variable in a model. To mask the information of a variable during validation, instead of removing the variable from the dataset, the PI method replaced it with random noise from other participants by shuffling the values of the variable (28). The relative importance of a variable was calculated as the accuracy decrease of the variable relative to the range of the accuracy decreases of all the variables (29).

In the RF model, the GI indicates how often a particular variable was selected for a split and how large its overall discriminative value was for the classification problem under study (27).

### eMethods 2. The description of K-fold cross-validation method

The dataset was split into 5 partitions, which instantiated 5 identical model building and validation processes. The LR and RF models were built on 4 partitions while the predictive ability indices were calculated using the remaining partition. Finally, the average predictive ability indices were calculated over the 5 partitions (30).

The eFigure 1 is available at *The Journals of Gerontology, Series A: Biological Sciences and Medical Sciences* online.

## Supporting information

eFigure 1

## Data Availability

All data used during the study are available from the corresponding author by reasonable request.

## Acknowledgments

DD and YC developed the original idea and designed the approach. ZX and DD searched the literature. ZX, QZ, XL, WW collected data. YC and ZX analyzed data. ZX, DD, and YC wrote the manuscript. QZ, XL, and WW revised the manuscript.

