## Supplementary figures and images for "Poor odor identification predicts mortality risk in older adults without neurodegenerative diseases: the Shanghai Aging Study"

### eFigure 1

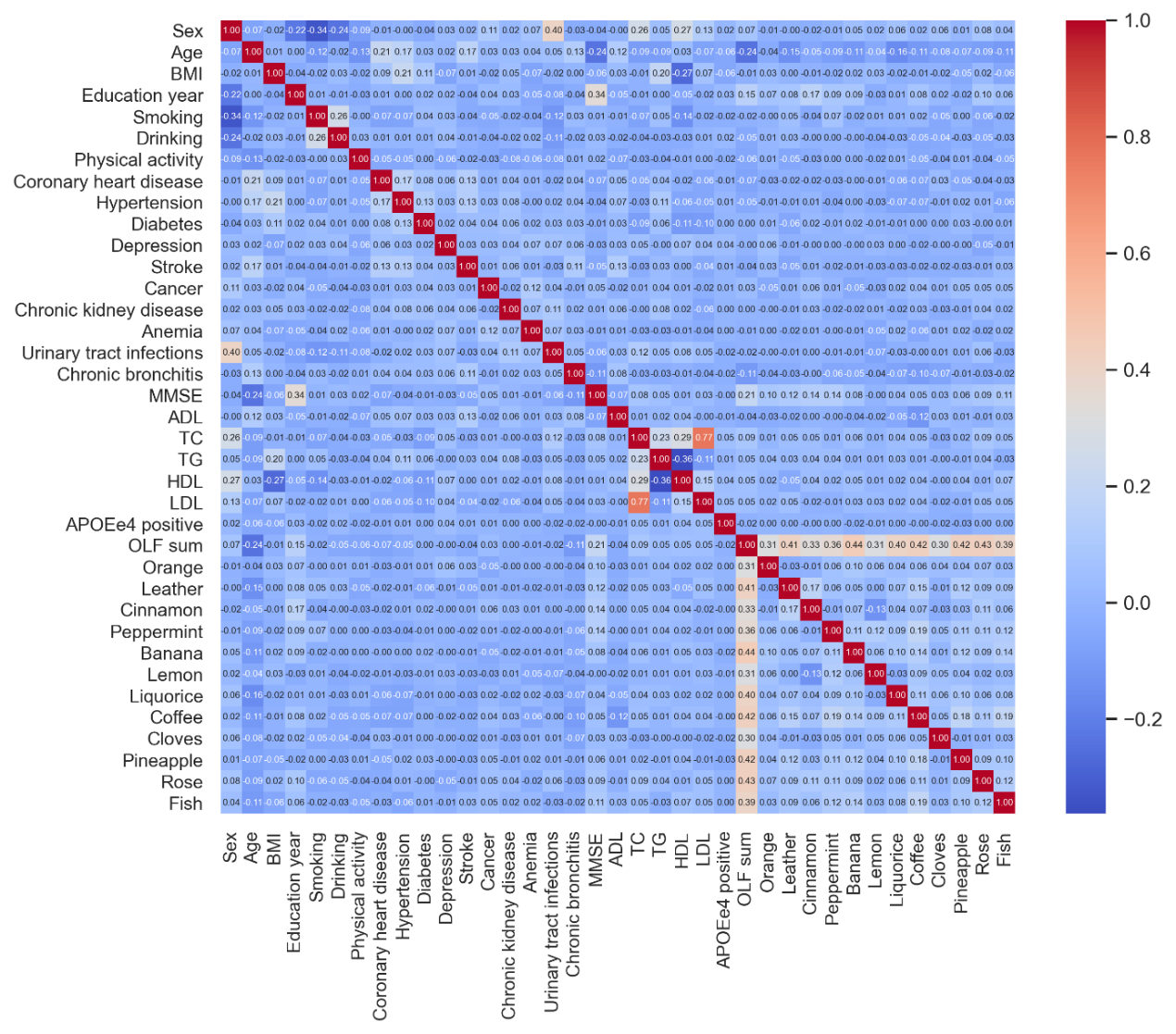

**eFigure 1. Correlation matrix heatmap of the predictor variables**
